# Body Image and Its Associated Factors among People Living with HIV: A Comprehensive Systematic Review and implications for integrated care

**DOI:** 10.1101/2025.05.04.25326771

**Authors:** Atena Pasha, Mohammad Jahanaray, Xiaoming Li, Shan Qiao

## Abstract

**Objectives:** People living with HIV (PLWH) face unique psychosocial challenges due to both infection and antiretroviral therapy (ART), one of which is body image disruption. Yet, a comprehensive synthesis of existing research on body image among PLWH is lacking. This study systematically reviewed relevant studies to explore body image issues, identify associated factors, and describe assessment methods and interventions targeting body image in this population.

**Methods:** Guided by the PRISMA, a thorough search of PsycINFO, PubMed, Embase, and Web of Science was conducted in January 2024, including empirical studies considering Body Image among PLWH published in peer-reviewed English journals, using search terms relevant to HIV and Body image. To include the latest articles, we conducted another round of searches in November 2024. NIH Study Quality Assessment Tools were used to assess the quality of the included studies, and a narrative synthesis was conducted to identify common themes, including definitions of body image, associated factors, measurement instruments, and interventions targeting body image among PLWH.

**Results:** From 2197 publications, 26 studies from 2004 to 2024 met the inclusion criteria, comprising a sample of 4095 PLWH aged 8 to 65 from different countries. Most of the studies were cross-sectional in design and varied in focus. Findings reveal that body image issues are prevalent among PLWH. The majority of studies demonstrated an association between negative body image and psychological comorbidities, including depression, anxiety, social withdrawal, and substance use. Body image dissatisfaction was also associated with physical health factors such as lipodystrophy. BMI measures reported in twelve studies indicated that BMI tends to increase with age in PLWH. Sixteen distinct body image measurement tools were used across studies. CBT-BISC was the only target intervention that showed effectiveness in mitigating body image disturbance and improving ART adherence among PLWH.

**Conclusion:** Body image issues represent a critical but often overlooked component of the biopsychosocial challenges faced by PLWH. This is the first comprehensive literature review to exclusively consider body image, associated factors, measurements, and target interventions among PLWH, which highlighted the need for comprehensive, culturally sensitive interventions that address both the physical and psychological dimensions of body image concerns.

## INTRODUCTION

According to the World Health Organization (WHO), HIV remains a major global public health issue, with an estimated 39.9 million people living with the virus by 2024 and approximately 42.3 million deaths. While there is currently no cure for HIV, access to effective prevention, diagnosis, treatment, and care has transformed HIV into a manageable chronic health condition, allowing people living with HIV (PLWH) to enjoy long and healthy lives [1]. However, one common challenge caused by HIV infection and Antiretroviral therapy (ART) treatment is the experience of side effects, especially changes in body image and composition [2].

Body image is a multidimensional construct conceptualized as an individual’s perceptions, thoughts, and feelings about their body or specific body parts [3]. The perception of body image among PLWH can undergo significant changes over time [4, 5]. Following diagnosis, individuals may experience a profound impact on their self-concept, feeling invaded by the virus and now living with a chronic condition [4]. As the infection progresses, other changes may occur, either caused by the virus itself or by the side effects of ART, requiring ongoing monitoring. Additionally, interactions with healthcare providers and social relationships may influence an individual’s self-image as a chronic patient [5]. The stigma associated with receiving a diagnosis of HIV can also result in a variety of detrimental emotional and physical outcomes. It might influence individuals’ perceptions of their own bodies, highlighting the significance of monitoring any alterations in body image that may arise during treatment [6–8].

Social, physiological, and psychological factors, including peers, age, gender, and culture, shape individuals’ perceptions of their bodies [9]. Changes in a person’s body due to illness or other factors can deeply impact their sense of self [10, 11]. Individuals with disabilities, chronic diseases, or changes in appearance are particularly impacted by the unique social context surrounding these conditions, which can lead to distinct reactions in social relationships and self-perception [12–14]. These changes can profoundly impact a person’s body image, encompassing their mental representation of their body identity, including self-perceptions and attitudes toward bodily appearance and function [9, 11].

Research among PLWH revealed that 79.5% of participants reported experiencing perceived bodily changes [15]. Data from the Women’s Interagency HIV Study revealed that self-reported fat gain and fat loss (either central or peripheral) were linked to a lower overall quality of life [16]. Moreover, studies indicate that an individual’s perception of their body image can significantly impact their adherence to ART medication among PLWH. Specifically, a negative body image has been correlated with non-adherence to medication [17] and a decrease in retention in care [18]. Other studies have demonstrated strong connections between HIV-related body changes and social isolation [19].

However, limited research has focused on understanding the psychological and social factors associated with body image concerns among PLWH. There is so far no comprehensive overview of body image issues related to HIV infection and associated factors. Additionally, there is no standardized measure to fully capture the concept of body image, although various methods, such as questionnaires and figure rating scales, have been used to assess body image. There is also a lack of interventions that address body image concerns among PLWH. Given the limited data on this topic and the demonstrated links between poor body image and negative clinical outcomes in PLWH, it is crucial to conduct a systematic review of existing literature regarding body image, associated psychosocial factors, different body image measurement instruments used among PLWH, and interventions developed for PLWH.

The current study aims to systematically review existing evidence from quantitative studies and randomized controlled trials (RCTs) on body image among PLWH. This is the first study of its kind to be conducted exclusively with PLWH that considered assessment tools and psychological interventions, which can be critically important for directing the distribution of resources and implementing various health programs to address physical, psychological, and social aspects of care for this population.

## METHODS

### Study design

This systematic review adhered to the preferred items for systematic reviews (PRISMA 2020) [20], ensuring the study’s credibility and reliability. The protocol was peer-reviewed and pre-registered on the Prospero website with ID number CRD42024530076. The search methodology and related steps are presented below.

### Search Strategy

In January 2024, a comprehensive search across multiple electronic databases was conducted, including PsycINFO, PubMed, Embase, and Web of Science. To include the latest articles, we conducted another round of searches in November 2024. This rigorous search strategy, aimed at selecting the most relevant and eligible publications, was initially based on preselected keywords and phrases. The keywords were a series of combinations of Body Image (e.g., “Body Dissatisfaction,” “Body Image Disturbance,” “Body Concern,” “Body Satisfaction”) and HIV (e.g., “HIV,” “AIDS,” “HIV/AIDS.”) (see **Supplementary Material**)

### Eligibility criteria

After conducting searches on PsycINFO, PubMed, Embase, and Web of Science, the articles were assessed for eligibility based on the following criteria: (1) They should be empirical studies in English and published in peer-reviewed journals; (2) They should focus on body image in PLWH; (3) They should use either validated or non-validated measures of one or more dimensions of body image. Additionally, there were no limitations on the age, gender, or ethnicity of the study participants.

The study excluded the following publications in accordance with its objectives: (1) studies published in a language other than English; (2) studies that only contain qualitative studies, commentaries, study protocols, literature reviews, conceptual papers, and conference abstracts.

### Information Sources

A two-part search strategy was used to identify studies that were eligible for inclusion. First, we searched electronic bibliographic databases for published studies using a string of preselected keywords and phrases. Second, we searched the reference lists of records that met the criteria for inclusion in the review and the reference lists of relevant, previously published review articles for other records that may be eligible for inclusion. The Rayyan cloud-based platform is used to manage the review process from the initial screening of records through to the selection of studies that are eligible for inclusion.

### Time period

All eligible studies published from 1 January 2000 to 24 November 2024 were included. The period from early 2000 to the present marks an era of large-scale ART coverage.

### Quality Assessment

The risk of bias/methodological quality of our included articles was assessed by two independent coders. The third researcher, who has experience in systematic and literature reviews, addressed the disagreements between the two. NIH Study Quality Assessment Tools were used to assess the quality of the included studies [21]. The Quality Assessment Tool for Observational Cohort and Cross-Sectional Studies was used for cross-sectional studies. Meanwhile, the Quality Assessment of Controlled Intervention Studies was used for Randomized Controlled Trial (RCT) studies.

Each question/dimension of the NIH tools was labeled with the following: “Not Reported (NR)” option used for studies that could potentially be answered “Yes” but the study did not report the required information; for instance, question 3 in the cross-sectional tool asks about the participation rate of eligible persons if a study only reports the number of participants but not the number of all of those who were approached initially this question would be “NR”. The “Cannot Determine (CD)” option is for those questions that are hard to answer, whether the study addresses them or not, because of lack of clarity. The “Not Applicable (NA)” option was used when one of the instrument’s criteria could not be evaluated due to the type of study (such as a cross-sectional design). Each study was labeled as each of the following:” Poor,” “Fair,” and “Good.”

### Data extraction and analysis

Data extraction was carried out using a standardized data extraction form designed to capture key study characteristics and findings. For each included study, the following information was extracted: author(s), year of publication, study design, sample size, population characteristics (e.g., age, gender, occupation), the body image definition and measurement tools used, key findings and associated factors related to body image, any reported interventions aimed improving body image among PLWH, limitation and future suggestion of included studies.

Two independent researchers extracted data from the included studies. All extracted data were recorded in a shared database to ensure consistency and accuracy. Any discrepancies in data extraction were resolved through discussion among the researchers.

### Data Synthesis

A narrative synthesis was conducted to identify common themes, including definitions of body image, associated factors, measurement instruments, and interventions targeting body image among PLWH. These themes informed the analysis by organizing and interpreting the extracted data, addressing the review’s research questions, and providing insights into the factors associated with body image. Additionally, the synthesis guided the categorization of measurement instruments and interventions, facilitating a comprehensive understanding of the methodologies and practices reported in the studies.

## RESULTS

### Quality Assessment

**Table 1** shows the quality assessment in this review. In the quality assessment of observational cohort and cross-sectional studies, several failed to meet specific criteria, impacting their overall robustness and validity. Notably, 16 studies (approximately 61.5%) did not report or deemed inapplicable the criteria related to participation rates of eligible persons (Q3), and three studies failed to have clear inclusion/exclusion criteria and uniformity in recruitment from similar populations (Q4). Similarly, 18 studies lacked sample size justification or discussion on statistical power (Q5), while 20 studies failed to measure exposures prior to outcomes (Q6). Based on the NIH tool description, some studies, like cross-sectional studies, would get “0” because of their study design type. Blinding of outcome assessors and loss to follow-up protocols were also inadequately reported or not applicable in all studies and 22 studies, respectively (Q12 and Q13). Lastly, seven studies did not adequately measure and adjust for key confounders (Q14), which is critical for isolating the exposure effects from other variables. It is important to note that most of our studies assessed the relationship between their variables of interest at a singular point in time. Consequently, responses to questions 6, 7, 10, 12, and 13 for nearly all these studies would correspondingly be “No” or “Not Applicable.” Therefore, the overall quality assessment of these studies was conducted based on eligible questions, intentionally excluding the non-eligible questions from the comprehensive evaluation.

**Table 1.** Quality Assessment of Included Studies.

| Authors | Q1 | Q2 | Q3 | Q4 | Q5 | Q6 | Q7 | Q8 | Q9 | Q10 | Q11 | Q12 | Q13 | Q14 | Total Score | Decision |
| --- | --- | --- | --- | --- | --- | --- | --- | --- | --- | --- | --- | --- | --- | --- | --- | --- |
| Blashill et al. (2011) | Y | Y | NR | NR | N | NA | NA | Y | Y | NA | Y | NA | NA | N | 5 | Fair |
| Lima et al. (2018) | Y | Y | Y | Y | Y | N | N | Y | Y | NA | Y | NA | NA | Y | 9 | Good |
| Fingeret et al. (2007) | Y | Y | Y | Y | N | Y | Y | Y | Y | N | Y | NA | Y | Y | 11 | Good |
| Junior et al. (2021) | Y | Y | NR | Y | Y | N | N | Y | Y | NA | Y | NA | NA | Y | 8 | Good |
| Corless et al. (2004) | Y | Y | NR | Y | N | N | N | Y | Y | NA | Y | NA | NA | N | 6 | fair |
| Lamb et al. (2018) | Y | Y | Y | Y | Y | Y | NR | NR | Y | Y | Y | NR | Y | CD | 10 | good |
| Gholizadeh et al. (2018). | Y | Y | NR | Y | N | N | N | NA | Y | NA | Y | NA | NA | Y | 6 | good |
| Blashill et al. (2010) | Y | Y | NR | N | N | N | N | Y | Y | NA | Y | NA | NA | Y | 6 | good |
| C. Martins et al. (2020) | Y | Y | NR | Y | Y | N | N | Y | Y | NA | Y | NA | NA | Y | 8 | good |
| Raggio et al. (2020) | Y | Y | NR | Y | N | N | N | Y | Y | NA | Y | NA | NA | Y | 7 | good |
| Klimek et al. (2020) | Y | Y | NR | Y | Y | Y | Y | Y | Y | Y | Y | CD | Y | Y | 12 | good |
| Blashill et al. (2017) | Y | Y | Y | Y | Y | Y | Y | NR | Y | Y | Y | NR | Y | Y | 12 | good |
| Blashill et al. (2014) | Y | Y | NR | Y | N | N | N | Y | Y | NA | Y | NA | NA | Y | 7 | good |
| Zanlorenzi et al. (2022) | Y | Y | Y | Y | Y | N | N | Y | Y | N | Y | NA | NA | Y | 9 | good |
| Huang et al. (2006) | Y | Y | Y | Y | N | N | N | N | Y | N | Y | NA | NA | Y | 6 | good |
| Guaraldi et al. (2008) | Y | Y | Y | Y | N | N | NA | N | Y | NA | Y | NA | NA | N | 6 | fair |
| Luzi et al. (2009) | Y | Y | Y | Y | N | N | N | Y | Y | NA | Y | NA | NA | Y | 8 | good |
| M. Martins et al. (2020) | Y | Y | NR | Y | Y | NA | NA | NA | Y | NA | Y | NA | NA | NA | 6 | good |
| Palmer et al. (2011) | Y | Y | NR | Y | N | Y | Y | Y | Y | CD | Y | NR | NA | Y | 9 | good |
| Sackey et al. (2019) | Y | Y | N | Y | N | N | N | Y | Y | NA | Y | NA | NA | Y | 7 | fair |
| Sharma et al. (2007) | Y | Y | NR | Y | N | NA | NA | Y | Y | NA | Y | NA | NA | Y | 7 | good |
| Martinez et al. (2005) | Y | Y | NR | Y | N | Y | NA | N | Y | Y | Y | NA | NA | Y | 8 | fair |
| Theodore et al. (2011) | Y | Y | NR | Y | N | N | N | Y | Y | CD | Y | NA | NR | Y | 7 | fair |
| Wilkins et al. (2016) | Y | Y | Y | Y | N | N | N | Y | Y | NA | Y | NA | NA | NA | 7 | good |
| Wu (2016) | Y | Y | NR | Y | N | N | N | NA | Y | NA | Y | NA | NA | NA | 5 | good |
| Soares et al. (2024) | Y | Y | Y | N | N | N | N | Y | Y | NA | NA | NA | NA | Y | 6 | good |

### Characteristics of Included Studies

As shown in **Table 2**, 26 studies out of 2197 publications included which were conducted between 2004 and 2024, with the majority of the studies carried out in the United States (17 studies) and Brazil (6 studies), and others were carried out in Italy (2 studies) and Canada (1 study). The sample sizes across these studies ranged from as few as 10 participants in Sackey et al. (2019) to 550 participants in Sharma et al. (2007); however, on average, the majority of the studies had around 150 participants. Recruitment sites include internet-based platforms such as discussion boards and listservs, outpatient clinics, primary care clinics, community health centers, general and specialized hospitals, and other settings like AIDS service organizations and private offices. The majority of the studies were cross-sectional (15 of the 26 reviewed studies). Three studies employed secondary analyses [22–24], while RCTs [25, 26], psychometric study [7, 27], and mixed-methods approaches [28, 29] were less common, each represented by two studies. Other study methods used include a correlation analysis [30] and a survey study [31].

**Table 2.** Sample characteristics of included papers.

| ID | Citation | Country | Design | Sample Size | Sample Characteristics | Recruitment site | Mode of HIV acquisition | Time of HIV diagnosis | Medication use | Comorbid Condition | BMI |
| --- | --- | --- | --- | --- | --- | --- | --- | --- | --- | --- | --- |
| 1 | Blashill & Vander wal(2011) | United States | Survey | 254 gay men with HIV | Age: 42.27 ±13.38; man;85.4% Caucasian,5.1% Hispanic/Latino, 4.3% Black/African, 2.4% Asian, 1.6% Multiethnic, 0.8% Native American, 0.4% Hawaiian Native | Various gay and HIV/AIDS-related Internet-based discussion boards and listserves | Not Reported | Most diagnosed with HIV more than 10 years ago (n = 78, 56.1%), 6 to 10 years (n = 20, 14.4%), 2 to 6 years (n = 28, 20.2%), or less than 1 to 2 years (n = 13, 9.3%). | 123 (88.5%) were taking medication to treat their conditions at the time of the study. | Not Reported | 26.37±6.34 |
| 2 | Lima et al. (2018) | Brazil | Cross-sectional | 111 PLWH | Age: 13 (10-15 years). | Outpatient clinic | Mother-to-child transmission | Not Reported | Not Reported | Not Reported | 18.52±2.62 |
| 3 | Fingeret et al. (2007) | United States | Secondary analysis | 77 People living with HIV/AIDS (PLWHA) | Age: 43.48 +_ 7.77 (27-64); 60 men, 17 women; 75% African American, 20% Caucasian, 5% Hispanic | Primary care clinic within a large, inner-city HIV/AIDS center | 32.5% male-to-male sexual contact, 39% heterosexual contact, 22% injection drug use, 6.5% other | Not Reported | Not Reported | Not Reported | Not Reported |
| 4 | Junior et al. (2021) | Brazil | Cross-sectional | 65 HIV-children and adolescents | Age: 12.2 (8-15 years). 30 Male and 35 Female | Federal University of Santa Catarina | Mother-to-child transmission | Not Reported | 83.1% were using ART | Not Reported | 17.92±2.65 |
| 5 | Corless et al. (2004) | United States | Correlational study | 40 HIV-patients | Age: 39.5 23 Male, 17 Female | Primary care clinic | Not Reported | Men = 9.2 years, Women = 7.2 years | Not Reported | Not Reported | Not Reported |
| 6 | Lamb et al. 2018) | United States | Randomized controlled trial (RCT) | 44 sexual minority men living with HIV | Age: 46 (SD=11); man; 64% White, 34% African American/Black, 23% Hispanic/Latino | Community health center in Boston | Not Reported | Not Reported | All taking ART | 42% with at least one psychiatric condition, 73% with two or more, 68% BDD, 46% MDD, 34% GAD | 27± 4.9 |
| 7 | Gholizadeh et al. (2018) | United States | cross-sectional | 105 sexual minority men living with HIV who reported having sex with a man within the previous 12 months. | Age: mean of 37 with a range of 18-65 years | General Hospital Infectious Disease Clinic and a community health center | Not Reported | 13.15 years (±8.2) | All taking ART | Not Reported | 27.96±4.67 |
| 8 | Blashill & Vander Wal (2010) | United States | cross-sectional | 124 HIV-positive gay and bisexual men | Age: 47.94 (SD=9.51); man; 85.5% Caucasian, 7.3% Hispanic/Latino, 4.8% Black/African American, 0.8% Multiethnic, 0.8% Asian, 0.80% Native-American | Various HIV/AIDS related Internet-based discussion boards and listservs | Not Reported | 1 year (1.6%)<br>1–2 years (4.0%)<br>2–4 years (10.5%)<br>4–6 years (8.9%)<br>6–8 years (11.3%)<br>8–10 years (4.8%)<br>10 years (58.9%) | HAART non-adherence for 10.57 years ± 1.72 for all participants | Not Reported | 25.60 ± 5 |
| 9 | C. Martins et al. (2020) | Brazil | cross-sectional study | 412 PLWHA | Age: 45.2 (SD=16.3); 60.8% women, 39.2% male | a follow-up service (STD/AIDS Specialized Service-SAE) | Not Reported | Not Reported | 90.20% used HAART over the past year | Not Reported | Not Reported |
| 10 | Raggio et al. (2020) | United States | Cross Sectional | 63 women living with HIV | Age: 51 (45-79); women; 58.7% African American/Black, 24% Caucasian, 2% American Indian/Alaska Native, 16% other, 11% Hispanic/Latina | a private office in the principal investigator's hospital | Not Reported | 14.1 ± 7.3 | 95.10% used ART at the time of survey completion | 70% depression, 11% anxiety | Not Reported |
| 11 | Klimek et al. (2020) | United States | Secondary data analysis | 22 sexual minority men with HIV and body image disturbance (43 gay, 1 bisexual) | Age: 46 (SD=11) (18-65 years); 63.6% White, 34.1% Black, 4.5% Native American, 4.5% other | Fenway Health, a community health center in Boston | Not Reported | Not Reported | Used ART for the past two months | 68.2% BDD, 18.2% ED, 95.5% at least one psychiatric diagnosis | Not Reported |
| 12 | Blashill et al. (2017) | United States | RCT | 44 sexual minority men living with HIV who reported elevated appearance concerns | Age: 46.18 (SD=11.03) (18-65); man; 34.10% African American/Black, 63.60% White, 4.5% Native American, 4.5% other, 22.7% Hispanic/Latino | Fenway Health, a community health center | Not Reported | Not Reported | Used ART for the past two months | 95.5% at least one psychiatric diagnosis, 72.72% two or more diagnoses, 68.20% Body dysmorphic disorder, 45.50% Major depressive disorder, 34.10% Generalized anxiety disorder, 27.30% Dysthymic disorder, 20.50% Substance dependence, 18.20% eating disorder, 15.90% Social anxiety disorder, 15.90% Alcohol dependence | 27.3±4.88 |
| 13 | Blashill et al. (2014) | United States | Cross Sectional | 106 gay and bisexual men living with HIV | Age: 47.5 (SD=7.8) (18-65); man; 12% Hispanic/Latino, 73% White, 23% Black/African American, 3% Native American, 4% other, 1% Asian | General Hospital Infectious Disease Clinic, and Fenway Health, a community health center | Not Reported | 13.1 ± 8.2 | Used ART for the past two months | Depressive symptoms (mean CES-D ≈ 20); lipodystrophy symptoms | 27.1±4.6 |
| 14 | Zanlorenzi et al. (2022) | Brazil | Cross-sectional | 60 children and adolescents diagnosed with HIV | Age: 11.93 (SD=6.99) (8-15 years); 32 females; 28 males | Outpatient clinic of a regional HIV reference center | Mother-to-child transmission | Not Reported | 83.33% were using ART | Lipoatrophy | 17.8±2.71 |
| 15 | Huang et al. (2006) | United States | Correlational study | 110 HIV-infected men | Age: 44 (38-50); man; 26% Hispanic, 8% Black, 59% White, 1% Asian, 6% other | an academic multidisciplinary adult HIV clinic | Not Reported | Not Reported | ART | 43.6%-48.2% Lipodystrophy (self-reported vs. physician-rated); 26.4% depression and anxiety, 65.5% AIDS | Not Reported |
| 16 | Guaraldi et al. (2006) | Italy | Cross sectional | 330 PLWH | Age: 41.5 (SD=6.5); 26.74% female, 73.25% male | HIV outpatient clinic | Not Reported | Not Reported | All were on ART for at least 6 months | 89.5% lipodystrophy | 22.5±3 |
| 17 | Luzi et al. (2009) | Italy | Cross sectional | 185 HIV-infected women | Age: 42 (SD=5); Females; | Routine HIV facility care | Not Reported | 16 years for FSD and 13.8 years without FSD | 86% are currently on ART at baseline | 81% lipodystrophy, 6% diabetes, 11% menopause | Not reported |
| 18 | M. Martins et al. (2020) | Brazil | Psychometric Study | 400 Brazilian people living with HIV/AIDS | Age: 41.12 (SD=11.68) (18-78); 67.8% male, 35% female; 53.8% Caucasian, 9.3% Black, 43% other | An HIV/AIDS ambulatory | Not Reported | Not reported | All were on ART | 51.8% Lipodystrophy | Not Reported |
| 19 | Palmer et al. (2011) | Canada | Secondary Analysis | 451 PLWH | Age: mean of 46. 24% female, 76% male; 33% Aboriginal, 67% Non-aboriginal | Centre for Excellence in HIV/AIDS (BCCfE) | Not Reported | Not Reported | 90% were currently on ART | 57% had depressive symptoms | Not Assessed |
| 20 | Sackey et al. (2019) | United States | Mixed-Method | 10 PLWH | Age: 54.95 (SD=3.02); 72.5% Black, non-Hispanic, 12.5% White, non-Hispanic, 10% Hispanic/Latino, 2.5% Mixed, non-Hispanic | Gay Men's Health Crisis (GMHC), a leading AIDS service organization | Not reported | Minimum years living with HIV was 18 years | 93% were on ART | 55% High cholesterol, 32.5% High blood pressure, 10% Type 2 diabetes, 20% Lipodystrophy, 27.5% Osteoporosis, 20% Opportunistic infection in last 5 years, 32.5% Arthritis, 20% CVD | 25.25±3.7 |
| 21 | Sharma et al. (2007) | United States | Cross-sectional | 550 (322 HIV-infected) older men with or at risk for HIV infection | Age: 55.2 (SD=4.5) (49-74); 12.4% White, 59% Black, 22.7% Hispanic, 5.9% other | Montefiore Medical Center | Not reported | 11 years (7-15) | 86.3% on ART | 43.8% Depressive Symptoms: of HIV-infected men, 36% Erectile Dysfunction | Not Assessed |
| 22 | Martinez et al. (2005) | United States | Cross-sectional exploratory study | 150 HIV-positive men and women | Age: 42 (22-65); 78.9% male, 21.1% female; 51.7% Caucasian, 21.9% Latino, 13.9% African American, 2.6% Asian, 6.6% other | Outpatient clinic (a public hospital HIV clinic and a university hospital HIV clinic) | Not Reported | 1 months-19 years | Not Reported | Not Required | Not Reported |
| 23 | Theodore et al. (2011) | United States | Cross-sectional | 42 HIV-positive Gay Bisexual Men | Age: 37.5 (SD=5.4); 47.61% White/Non-Hispanic, 40.47% Latino/Hispanic, 9.52% African American, 2.38% other | Recruited at a "gay beach" in South Florida | Not Reported | Not Reported | 30 participants (71.4%) used HIV medication | Not Reported | Not Assessed |
| 24 | Wilkins et al. (2016) | United States | Cross Sectional | 119 Youths with HIV | Age: 20.7 (SD=1.98); 89 Male, 30 female. Caucasian: 5, Hispanic: 1, African American: 113 | Infectious diseases clinic | 83.21% behavioral, 16.78% perinatal | 24.75 ± 5.36 | 110 participants (76.9%) were on ARVs | Not Reported | 25.3±6.1 |
| 25 | Wu et al.<br>(2016) | United States | Mixed-Method | 96 adults living with HIV | Age: 48.8 (SD=8.3).<br>84 Male, 12 Female.<br>Caucasian: 64, Asian: 1,<br>Hispanic: 7, African American: 20 | HIV Clinic | Not reported | Not reported | All had a history of ARV therapy | Facial lipoatrophy | Not Assessed |
| 26 | Soares et al.<br>(2024) | Brazil | Cross-sectional | 231 adults living with HIV | Age: 40 (SD=8.9).<br>154 Male, 77 Female | Immunodeficiencies<br>Outpatient Clinic | Not Reported | Mean of 42 months. | 72.3% were on ART, with 28.5% exposed to protease inhibitors (PIs) | 40% had self-reported lipodystrophy | Not Assessed |

### Sociodemographic Characteristics of PLWH

**Table 2** summarizes the sociodemographic characteristics of PLWH. Participants’ ages ranged from 8 to 65. Studies with the adult group (aged 18 and above) had a mean age of 44.6 years across the studies, with a range of mean ages from 37 to 55.2. For the non-adult group (aged under 18) the mean age across studies was 15.1 years, with a range of mean ages from 11.93 to 20.7 years. Zanlorenci et al. [32] reported the lowest mean age (11.93 ± 6.99), and Sharma et al [33] reported the highest mean age (55.2 ± 4.5). Out of a total of 4,095 PLWH, 3062 were males, and 1033 were females. Gender distribution across the studies predominantly featured male participants, although several studies included substantial female representation [8, 32, 34–36]. However, 11 studies focused on only male participants [18, 23, 25, 26, 28, 31, 33, 37–40]. The racial composition of participants varied across studies, with a significant proportion of participants identifying as Caucasian (1149) and Black/African American (702), Hispanic/Latino (243), Asian (12), Native-American (10), Multiethnic (5), Native Hawaiian (1), and 232 categorized as Other.

The modes of HIV acquisition were not reported in most of the included articles, except for five studies, where four out of them mentioned Mother-to-child transmission [32, 34, 35, 41], and Fingeret et al. [22] reported that 32.5% acquired HIV through male-to-male sexual contact, 39% through heterosexual contact, and 22% via injection drug use. The duration since HIV diagnosis varied considerably among the studies. A notable portion of participants (56.1%) had been diagnosed with HIV for over a decade [28, 33, 36, 38, 40, 42, 43]. Other studies consist of participants with shorter durations of diagnoses, with groups diagnosed between 6 to 10 years (14.4%), 2 to 6 years (20.2%), and less than 1 to 2 years (9.3%).[18, 30, 31]⍰. The duration of diagnosis also varied by gender, with men having an average of 9.2 years and women 7.2 years.

Only 8 studies reported the type of medications or treatment that PLWH were on at the time of the study. different categories of ART were most commonly reported, such as Nucleoside Reverse Transcriptase Inhibitors (NRTIs), Non-Nucleoside Reverse Transcriptase Inhibitors (NNRTIs), and Protease Inhibitors (PIs) [28, 30, 34, 35, 41–43].

### Body Image Dimensions and Measurement

The studies have focused on different body image concentrations. Fingeret et al. [22], Blashill and Vander Wal [18], Blashill and Vander Wal [31], Theodore et al. [39], and Gholizadeh et al. [40] concentrated on PLWH body dissatisfaction. While Blashill et al. [38], Blashill et al. [26], Lamb et al. [25], and Klimek et al. [23] delved into body image disturbance and its nuances in PLWH. Martinez et al. [27], Guaraldi et al. [44], Wilkins et al. [42], and Zanlorenci et al. [32] studied body image perception among PLWH. Luzi et al. [43], Raggio et al. [36], and Soares et al. [41] focused on PLWH body satisfaction. Negative body image among PLWH was assessed by Sharma et al. [33] and Palmer et al. [24]. Corless et al. [30], Huang et al. [37], Lima et al. [34], and Junior et al. [35] aimed at assessing overall body image. Other studies by Wu et al. [29], Sackey et al. [28], M. Martins. et al. [7], and C. Martins. et al. [8] explored Cognitive and Behavioral Aspects of PLWH Body Image Anxiety, Body Image Attitude, Body Image Distress and Discomfort, and self-perception of body image, respectively.

Among the 26 included studies, 16 different body image measures were utilized to assess body dissatisfaction, perception, and related psychological constructs among PLWH (see **Table 3**). The reliability of these measures, reported as Cronbach’s Alpha, varied across studies, indicating differences in internal consistency. The Assessment of Body Change Distress scale (ABCD) was one of the most frequently used instruments, appearing in Guaraldi et al. [44] (α = 0.94), Blashill et al. [38] (α = 0.84), and Wu et al. [29] (α = 0.98), also in Luzi et al. [43] and Raggio et al., [36] but with no reports on the internal consistencies. This scale demonstrated high reliability, making it a robust tool for evaluating body disturbance and satisfaction associated with ART-related physical changes. The Multidimensional Body-Self Relations Questionnaire (MBSRQ) was used in Blashill and Vander Wal [31] (α = 0.92) and Wilkins et al. [42] (α = 0.74). These findings suggest that the MBSRQ remains a reliable tool for assessing body image dissatisfaction and perception among PLWH.

**Table 3.** Body image measurement instruments and key findings.

| ID | Citation | Name of instrument | Instrument focus | Cronbach's alpha | Findings |
| --- | --- | --- | --- | --- | --- |
| 1 | Blashill & Vander Wal (2011) | MBSRQ | Body Dissatisfaction | Appearance Evaluation: 0.92<br>Appearance Orientation: 0.88<br>Fitness/Health Evaluation: 0.83<br>Fitness/Health Orientation: .092<br>Illness Orientation: $\alpha = 0.76$ | Men in the AIDS group reported being less fit and in poorer health than HIV positive and HIV negative men. Surprisingly, there were no significant differences between groups on appearance evaluation. |
| 2 | Lima et al. (2018) | The silhouette scale | Body image | Not Reported | Body Image dissatisfaction was linked to a desire to increase body size (38.6%), especially in males, and was associated with lower anthropometric measures like skinfolds and UAMA (Upper Arm Muscle Area), while gender, age, weight, BMI, and UAMA explained 42% of body image score variation. |
| 3 | Fingeret et al. (2007) | BIS | Body image/appearance dissatisfaction | 0.88 | In PLWHA, body image concerns were associated with depression, anxiety, stress, and lower social support, yet smoking cessation rates were highest at moderate concern levels and lowest at both high and low extremes, suggesting a unique curvilinear relationship not solely tied to weight concerns. |
| 4 | Junior et al. (2021) | Scale of body silhouettes | Body image | Not Reported | In HIV-infected children and adolescents, body image dissatisfaction was prevalent, with 38.6% wanting to increase body size (notably males) and 40.7% of a control group wanting to reduce it. Among female PLWH, those desiring to reduce body weight had higher BMI and fat mass compared to those satisfied or wanting to increase it, while no significant differences were found in males across body image categories. |
| 5 | Corless et al. (2004) | MOS-HIV | Body image (actual and ideal) | 0.90 | It found that weight change in men was related to better mental health, vitality, and quality of life scores. For women, weight change did not significantly correlate with these domains. Body image scores were higher for women than men, and there was no significant difference in body image between participants with HIV and those with AIDS. |
| 6 | Lamb et al. (2018) | BDD-YBOCS | Body Image Disturbance | 0.84 at baseline to 0.93 at the 3-month follow-up | Participants assigned to CBT-BISC reported statistically significant reductions in body image disturbance post-intervention, which subsequently predicted changes in ART adherence from post-intervention to long-term follow-up. One pathway in which CBT-BISC positively impacts ART adherence is through reductions in body image disturbance. |
| 7 | Gholizadeh et al. (2018) | The Muscle Dysmorphic Disorder Inventory (MDDI), Appearance Orientation subscale of the Multidimensional Body-Self Relations | Body Dissatisfaction | 0.72 | A generalized linear model identified a significant interaction ($b = 0.08$ [95% CI: 0.01, 0.16], $p = 0.033$) such that when appearance investment was low, body dissatisfaction was associated with fewer condomless anal sex acts; when appearance investment was high, body dissatisfaction was associated with increased condomless anal sex. |
| 8 | Blashill & Vander Wal (2010) | MBSRQ | Body Dissatisfaction | 0.91 | The results of the moderated mediation models indicated that depression mediated the relationship between body dissatisfaction and HAART non-adherence for men with high levels of body dissatisfaction but not for those with moderate or low levels. Furthermore, depression also mediated the relationship between body dissatisfaction and HAART non-adherence among men who self-reported an AIDS diagnosis. |
| 9 | Martins et al. (2020) | Silhouette scales | The self-perception of body image (SPBI) | Not Reported | Patients who use HAART are experiencing new challenges, such as metabolic and morphological body changes, which may affect self-perceived BI. Based on the results obtained, it can be concluded that NSPBI in HIV-infected patients was associated with Nadir CD4, variations in BMI, particularly LW and obesity, self-perception of specific body changes and mental health (higher prevalence of depressive symptoms). |
| 10 | Raggio et al. (2020) | The AIDS Clinical Trials Group Assessment of Body Change and Distress scale | Body Satisfaction | Not Reported | Findings present a clear profile of the psychosocial and body image challenges associated with prevalent lipodystrophy among aging, actively treated WLWH. Our data reveal persistent body image disturbance among patients well over a decade after being diagnosed with HIV. Also, social support was not significantly associated with lipodystrophy severity or either of the prevalent lipodystrophy phenotypes in bivariate analyses. More than a third of women reported severe/diffuse lipodystrophy, defined herein as three or more body parts evidencing fat redistribution. |
| 11 | Klimek et al. (2020) | BIDQ, BICSI | Body Image Disturbance, Body Image Coping Skills | 0.88 at baseline and 0.96 at follow-up | CBT-BISC significantly reduced body image disturbance and improved coping skills. Latent difference score mediation indicated that changes in acceptance and cognitive reappraisal significantly predicted body image disturbance changes. |
| 12 | Blashill et al. (2017) | BDD-YBOCS, the Brown Assessment of Beliefs Scale (BABS) | Body image disturbance | BDD-YBOCS: 0.84 at baseline and 0.93 at the 3-month follow-up<br>BIDQ: 0.88 at baseline and 0.96 at the 3-month follow-up. | The study demonstrates that CBT-BISC effectively reduces body image disturbance and depressive symptoms while improving adherence to HIV treatment among sexual minority men with HIV. These improvements are substantial and maintained over the long term, indicating that integrated therapeutic approaches that address both mental health and physical care are crucial for this population. |
| 13 | Blashill et al. (2014) | Body Change and Distress Questionnaire–Short Form (ABCD-SF), MBSRQ | Body Image Disturbance | 0.84 | The data fit the model well, with all theorized pathways being significant. Lipodystrophy severity and appearance orientation were associated with elevated body image disturbance. In turn, body image disturbance was related to poorer ART adherence and increased HIV sexual transmission risk behaviors, through the mechanisms of elevated depressive symptoms and poor condom use self-efficacy |
| 14 | Zanlorenci et al. (2022) | Silhouette scale | Body image perception | Not Reported | The results indicated that 53.13% of women and 53.57% of men were dissatisfied with their BI. Lower subscapular skinfold and higher calf skinfold values were associated with Body Image dissatisfaction in females. Pre-pubertal maturation stage, higher economic level, lower concentrations of CD4+ lymphocytes, lower viral load, lower level of physical activity, and longer time in front of the computer and/or video game were associated with Body Image dissatisfaction in males. |
| 15 | Huang et al. (2006) | BIQLI, SIBID-S | Body image | Not Reported | Compared to HIV-infected men who denied lipodystrophy, HIV-infected men with self-reported lipodystrophy demonstrated poor body image. Physician-rated lipodystrophy was significantly associated with both body image subscale scores. |
| 16 | Guaraldi et al. (2006) | ABCD | Body Image Perception | 0.94 | Preliminary evidence supports the reliability and validity of the Italian version of the ABCD in people with HIV and lipodystrophy. |
| 17 | Luzi et al. (2009) | Italian version of the ABCD | Body Satisfaction | Not Reported | Desire, arousal, and satisfaction domains were associated with interference of body changes with habits, social life, and attitudinal aspects of body image. Lubrication and orgasm domains of the Female Sexual Function Index (FSFI) were associated with body image satisfaction. Lipodystrophy's impact on body image was the sole risk factor associated with Female sexual dysfunction (FSD). This implies that changes in body composition and appearance, which can be significant in individuals with HIV undergoing treatment, play a critical role in influencing sexual function. |
| 18 | Martins et al. (2020) | Brazilian Portuguese version of DAS-24 | Body Image Distress and Discomfort | 0.94 | The Brazilian Portuguese version of DAS-24 seems to be a psychometrically sound scale for measuring body image distress for PLWHA. |
| 19 | Palmer et al. S. (2011) | BIS | Negative Body Image | Not Reported | Of 451 The Longitudinal Investigations into Supportive and Ancillary Health Services (LISA) participants, 47% reported negative body image. The adjusted multivariate analysis showed participants who reported high stigma in the presence of depressive symptoms were more likely to have negative body image compared to people reporting low stigma and no depressive symptoms (adjusted odds ratio [AOR]: 2.41, confidence interval [CI]: 1.244.68). The estimated probability of a person having a positive body image without stigma or depression was 68%. When stigma alone was included, the probability dropped to 59%, and when depression was included alone, the probability dropped to 34%. Depressive symptoms and high stigma combined resulted in a probability of reporting positive body image of 27%. |
| 20 | Sackey et al. (2019) | MBAS | Body Image Attitude | 0.79 | Participants reported challenges accessing healthy food and eating fruits and vegetables. Living with HIV/AIDS also influenced their food options, which in turn affected physical activity and body image. |
| 21 | Sharma et al. (2007) | Self-reported | Negative Body Image | Not Reported | 31% of participants reported negative body image, which was independently associated with increased BMI, self-rated fair/poor health, depression, and erectile dysfunction, but not HIV status. |
| 22 | Martinez et al. (2005) | BIS | Body Image Perception | 0.91 | The BIS has good construct validity and is a highly reproducible measure of self-perception of body image in HIV-infected people. |
| 23 | Theodore et al. (2011) | BSQ | Body Dissatisfaction | Not Reported | The study identified that body dissatisfaction explains a small but significant variance in methamphetamine use among HIV-positive gay and bisexual men after accounting for the effects of circuit party attendance and the use of alcohol and marijuana. This association highlights the psychological impact of body image issues on substance use behaviors within this demographic. |
| 24 | Wilkins et al. (2016) | MBSRQ, Figure Rating Scale (FRS) | Body Image Perception | 0.74 | Results showed a normative global body image on the MBSRQ-Appearance Scales. Some subscale elevations were observed, including decreased interest in self-care and appearance, as well as concerns with individual body areas. Overall, youth reported a preference for their own body shape on the Figure Rating Scale; however, 41 % of youth classified as “overweight” per CDC body mass index reported contentment with their current body size. Further, 47 % of youth classified as “normal” weight desired to have a larger body size. Youth identified as men who have sex with men most often reported desiring larger body size. |
| 25 | Wu et al. (2016) | ABCD, MOS-HIV | Cognitive and Behavioural Aspects of Body Image Anxiety | 0.98 | The study found that the Facial Appearance Inventory (FAI) is a reliable and valid tool for assessing the impact of ARV-associated facial lipoatrophy on quality of life in PLWH. The FAI correlated more strongly with mental health than physical health, indicating the significant psychological impact of facial lipoatrophy. More severe lipoatrophy was associated with worse FAI scores, underscoring its negative effect on quality of life. Additionally, individuals with less education and those with darker skin types reported less impairment, suggesting variations in the perceived impact of lipoatrophy across different demographic groups. |
| 26 | Soares et al. (2024) | Self-Reported | Body Image Satisfaction | Not Reported | The study found that 40% of participants experienced self-reported lipodystrophy, significantly affecting their body image, particularly through changes like fat loss in visible areas and increased waistlines. Notably, the use of lipid-lowering medications was positively correlated with improved self-perceptions of body image. Objective measurements via anthropometric and biochemical profiles were more effective than subjective reports, revealing that patients often perceived their body condition as worse than it was. |

The silhouette scale, commonly used to measure perceived body size, was employed in Lima et al. [34], Martins, C et al. (2020), Junior et al. [35], and Zanlorenci et al. [32], but none of the studies reported the internal consistency. The reliability of this the tool to ass body image perception among PLWH requires further exploration. The Body Image Scale (BIS), another used measure, was utilized in Martinez et al. [27] (α = 0.91) and Fingeret et al. [22] (α = 0.88), indicating strong internal consistency. Additionally, the Body Dysmorphic Disorder modification of the Yale-Brown Obsessive-Compulsive Scale (BDD-YBOCS), which specifically assesses body dysmorphic concerns, was employed in Blashill et al. [26] (α = 0.93) and Lamb et al. [25] (α = 0.93).

Several other body image scales were used in individual studies, demonstrating varied but generally high reliability. The Medical Outcomes Study-HIV (MOS-HIV) instrument used in Corless et al. [30] showed a reliability score of 0.90. The Situational Multidimensional Body-Self Relations (MBS), used by Blashill and Vander [18], exhibited a Cronbach’s Alpha of 0.91, while the Male Body Attitudes Scale (MBAS) employed by Sackey et al. [28] showed a reliability coefficient of 0.79. Lastly, the Body Image Disturbance Questionnaire (BIDQ) used in Klimek et al. [23] and the Body Image Quality of Life (BIQLI) and the Brazilian Portuguese version of the Derriford Appearance Scale 24 (DAS-24), used in M. Martins et al. [7], demonstrated high reliability (α = 0.96 and α=94, respectively).

### Body images, correlates, and consequences

As shown in **Table 3**, several studies reported high levels of body dissatisfaction and negative body image perceptions among PLWH related to physical changes induced by HIV and its treatment. Martinez et al. [27] found that HIV-positive individuals perceived their body image as significantly worse post-infection compared to pre-infection, a finding that may reflect the intersection of HIV-related stigma and body image ideals. Gholizadeh et al. [40] highlighted specific dimensions of body image, noting a mean appearance investment score of 3.61 and a body dissatisfaction score of 31.35, indicating considerable concerns related to appearance and dissatisfaction. Sharma et al. [33] differentiated between positive and negative body image, finding varying degrees of agreement and disagreement among a large sample of 550 male participants. On the other hand, Lamb et al. [25] observed significant reductions in body image concerns over time through a randomized controlled trial (CBT-BISC scores decreased from 25.36 at baseline to 8.24 at three months and 8.08 at six months).

BMI was commonly included as a correlate of body image in the literature. While BMI reflects body composition, body image is a perceptual and psychological construct, shaped by individual and sociocultural factors. Therefore, BMI does not directly predict body image outcomes, and associations between the two were often weak, inconsistent, or even contradictory. For instance, a study conducted among HIV-positive youth showed that those classified as overweight were often satisfied with their body size, while individuals with normal BMIs expressed a desire for a larger body (Wilkins et al., 2016). This divergence may be due to reliance on self-reported BMI in the majority of the included studies, a method prone to misreporting and social desirability bias. These inconsistencies suggest that while BMI may influence or reflect body image concerns among PLWH, it should be interpreted cautiously as it does not capture the subjective and multidimensional nature of body image.

Metabolic and morphological changes resulting from HIV and ART use have a profound impact on body image and overall well-being. Lipodystrophy is characterized by either partial or complete loss of fat in certain areas of the body, including the face and limbs, along with abnormal fat accumulation in other areas, such as the abdomen [45]. Lipoatrophy, on the other hand, specifically refers to the loss of subcutaneous fat, typically in the face and extremities, without the central fat accumulation seen in lipodystrophy [46]. Lipodystrophy and lipoatrophy both contribute to substantial psychological distress, social withdrawal, and a reluctance to disclose one’s HIV status due to fears of visible HIV-related stigma [44]. The psychosocial burden of these physical changes is particularly pronounced in individuals with facial lipoatrophy, as they often report lower self-esteem and higher rates of social anxiety [29]. Five studies reported the prevalence of lipodystrophy among participants, which ranged from 27.5% to 81%. Soares et al. [41] found that 40% of PLWH in their study reported self-perceived lipodystrophy, with significant correlations between lipodystrophy, elevated cholesterol, insulin resistance, and body image dissatisfaction, highlighting the metabolic complexities associated with ART use.

Several sociodemographic factors, including gender, minority status, and age influence the way PLWH perceive their bodies, with notable variations across different subgroups. Women living with HIV consistently report higher levels of body dissatisfaction compared to men. C. Martins, et al. [8] found that 74.3% of patients exhibited negative self-perception of body image, with higher prevalence among women (81.2%) compared to men (65.6%). Among men, sexual minority status emerged as a critical factor influencing body image concerns. Theodore et al. [39] found that HIV-positive gay and bisexual men exhibited high levels of body dissatisfaction, which was positively correlated with methamphetamine use. These findings suggest that body image concerns among sexual minority men may be linked to both internalized stigma and cultural expectations within LGBTQ+ communities.

Body image concerns fluctuate across different age groups among PLWH, particularly in response to lipodystrophy and ART-associated weight changes. Wilkins et al. [42] examined body image in adolescents living with HIV and found that youth displayed complex perceptions of body satisfaction. Notably, 41% of overweight youth were satisfied with their body size, whereas 47% of normal-weight youth desired a larger body size, highlighting age-specific patterns in body dissatisfaction. Conversely, studies focusing on older adults with HIV have reported heightened concerns related to weight gain, facial lipoatrophy, and visible signs of aging. Wu et al. [29] demonstrated that facial lipoatrophy was a significant factor impacting self-perceived body image and overall quality of life, reinforcing the psychosocial burden of ART-induced changes in body composition among older individuals. Studies also indicate racial and ethnic disparities in body image perception among PLWH. In this regard, Wilkins et al. [29] found that Black and Latino youth living with HIV reported distinct body image concerns, with men who have sex with men (MSM) expressing a stronger desire for a larger body size.

Psychiatric conditions, including depression, anxiety, and BDD, frequently co-occur with HIV and contribute to heightened body dissatisfaction. Studies by Lamb et al. [28], Raggio et al. [39], Klimek et al. [26], and Blashill et al. [29] documented high incidences of psychiatric conditions among PLWH, including body dysmorphic disorder (BDD), Major depressive disorder (MDD), generalized anxiety disorder (GAD), and substance use disorder (SUD). Notably, Lamb et al. [28] found that 95.5% of participants met the criteria for at least one psychiatric diagnosis, with 68.2% diagnosed with BDD. MDD was reported in 26.4% to 70% of participants, while anxiety disorders were noted in 11% to 34% of participants. The intersection of stigma, social exclusion, and depressive symptoms further exacerbates negative body image perceptions, with individuals experiencing both stigma and depression being more than twice as likely to report severe body dissatisfaction compared to those without these psychological stressors [24]. Several studies indicated an association between internalized stigma and indices of poor body image (e.g., Brewster et al. [47] and Hart et al. [48]). Studies such as Palmer et al. [24] reported strong associations between HIV-related stigma and body image concerns, particularly among individuals who experience depressive symptoms. Fingeret et al. [22] highlighted that the stigma surrounding physical changes due to HIV treatment often led to social withdrawal and heightened self-consciousness about appearance.

Body image may influence treatment adherence, mental health, and health-related quality of life among PLWH. For example, body image distress has been associated with poor adherence to ART, potentially leading to disease progression and a deteriorating self-perception of health [27]. Luzi et al. [43] reported that female sexual dysfunction (FSD) was directly linked to body image dissatisfaction. Similarly, among HIV-positive gay and bisexual men, body dissatisfaction has been associated with increased methamphetamine use, suggesting a pattern of self-medication to cope with body image distress [39]. Nutritional concerns further complicate these challenges; Wilkins et al. [42] found that while many HIV-positive youth classified as overweight were content with their body size, those with normal BMIs often desired a larger body, underscoring the unique psychosocial dimensions of body image in this population.

### Body image interventions

A key approach that stands out across a few studies is the use of Cognitive Behavioral Therapy for Body Image and Self-Care (CBT-BISC), a structured intervention designed to simultaneously address body image concerns and self-care behaviors, including ART adherence. Initially, all participants underwent Session 1 of Life-Steps, a single-session intervention that focuses on ART adherence and defining sexual health goals. Then, participants were assigned to either the Enhanced Treatment as Usual (ETAU) or CBT-BISC condition. In the ETAU condition, participants engaged in biweekly meetings with the project coordinator over the following three months. These sessions, lasting approximately 15 minutes, involved reviewing ART adherence via Wisepill data, addressing errors, and providing brief mental health referrals. Participants in the CBT-BISC condition received weekly individual sessions over three months (12 total sessions), with each session lasting around 50 minutes. CBT-BISC followed a structured manual, incorporating seven modules that targeted both body image and ART adherence. The first session was dedicated to orienting participants to the CBT framework and body image concepts. Subsequent modules integrated a range of cognitive behavioral techniques, such as mindfulness, perceptual retraining, cognitive restructuring, and exposure-based interventions to reduce body image disturbance. These strategies aimed to address negative body image and improve the participant’s overall well-being, which could, in turn, positively influence ART adherence. Sessions also included a review of adherence data, with body image discussions being framed around participants’ goals for ART adherence. The final module, relapse prevention, focused on reviewing learned skills and preparing participants for potential future challenges [26].

In the Lamb et al. [25] study, participants in the CBT-BISC group showed statistically significant reductions in body image disturbance, which were sustained at both three-month and six-month follow-ups. Notably, these reductions in body image concerns positively influenced ART adherence, highlighting a crucial link between body image and HIV treatment engagement. At six months post-treatment, 91% of participants in the CBT-BISC group maintained their improvements, compared to only 27% in the control group receiving enhanced treatment as usual (ETAU). Additionally, on-time ART adherence significantly improved (dppc^2^ ^1^= 0.94, p = .01), along with reductions in depressive symptoms (dppc^2^ = 1.17, p = .008) and increased global functioning (dppc^2^ = 3.39, p < .001).

Blashill et al. [26] further reinforced the effectiveness of CBT-BISC, showing that the intervention led to substantial reductions in body image disturbance, sustained ART adherence improvements, and significant decreases in depressive symptoms. Moreover, Klimek et al. [23] explored the specific skills-based mechanisms driving CBT-BISC efficacy, demonstrating that reductions in avoidance and appearance-fixing behaviors were significant contributors to body image improvement. However, their findings emphasized that acceptance and cognitive reappraisal strategies were the strongest mediators of long-term success (β = −0.47, p = .001), accounting for the most variance in body image improvements across conditions. This suggests that CBT-BISC is most effective when it enhances adaptive cognitive processing and reduces maladaptive body image-related coping strategies.

## DISCUSSION

This systematic review explored body image concerns and associated factors among PLWH. By analyzing 26 studies conducted between 2004 and 2024, key themes were identified across measurement tools, sociodemographic characteristics, biopsychosocial factors, and consequences, and intervention strategies. The findings underscore the multifaceted nature of body image issues in PLWH and reveal significant gaps in the literature that warrant further investigation.

### Key Findings

The findings reveal a high prevalence of body image dissatisfaction and disturbances among PLWH, such as a study conducted by C. Martins et al. [8] that reported rates of body dissatisfaction as high as 74.3%. Body image was associated with physical and clinical factors such as ART adherence and lipodystrophy [22, 24, 26, 28, 31, 32, 37, 38, 43, 44], psychological factors such as anxiety, depression, and stress [22–24, 33, 36, 38], and sociological factors, including stigma and social support [32, 38, 40, 43].

Physical changes, such as lipodystrophy and altered body composition, were prominent risk factors for developing negative body image among PLWH. These findings likely arise because lipodystrophy, a visible and stigmatized side effect of ART, directly affects appearance, leading to heightened self-awareness and dissatisfaction. BMI also emerged as a critical factor associated with body image. Studies like those by Junior et al. [35] noted that both lower and higher BMI values impacted body satisfaction, though through different mechanisms. Lower BMI was associated with perceptions of frailty and vulnerability to illness, while higher BMI raised concerns about stigma and societal ideals of body aesthetics.

Sociodemographic characteristics, including gender, age, and racial composition, appear to influence body image perceptions. The predominance of male participants in the reviewed studies reflects a gender bias in the existing literature. However, studies that included substantial female representation (e.g., Martins, C et al., [8]) reported that women with HIV exhibited higher rates of body dissatisfaction (81.2%) compared to men (65.6%). This discrepancy can be attributed to societal beauty standards that disproportionately emphasize physical appearance for women, amplifying the impact of HIV-related changes. This can also be explained by the negative effects of the mass media on body image perception, as body image dissatisfaction is strongly related to standards imposed by society and culture [49].

Furthermore, variations in racial and ethnic representation across studies suggest that cultural and contextual factors may play a role in shaping body image experiences, though this area remains underexplored. Junior et al. [35] noted that cultural norms surrounding body appearance and health perceptions could modulate body image concerns, particularly in settings with strong societal expectations around physical appearance. Such cultural pressures may exacerbate or mitigate body image concerns depending on the societal context and support structures available [50].

Social support and structured interventions acted as protective factors for body image issues among PLWH. Lamb et al. [25] reported that CBT-BISC not only improved body image perceptions but also enhanced positive coping mechanisms and adaptive emotion regulation strategies such as cognitive reappraisal and acceptance. Positive self-image and resilience were further bolstered by participation in structured interventions and access to affirming healthcare environments [25]. This suggests that supportive interventions can empower PLWH to challenge negative self-perceptions and build resilience against external judgments. Furthermore, social networks and community support emerged as critical buffers against the adverse effects of stigma and physical changes.

In conclusion, the reviewed studies suggest that addressing both risk and protective factors through targeted interventions could significantly mitigate body image concerns and improve overall well-being in PLWH.

### Knowledge Gaps

The majority of included studies were conducted in a limited number of countries, predominantly the United States, Brazil, Canada, and Italy, which limits the generalizability of findings to other regions, particularly low-income countries and non-Western settings. Our findings also showed that gender differences in body image dissatisfaction were often inadequately examined, with female participants experiencing higher levels of distress but receiving less research attention. In this regard, investigating how cultural and gender norms shape body image perceptions could lead to more targeted interventions for at-risk groups.

In seven studies, key confounders were not adequately measured or controlled. Factors such as HIV disease progression, medication side effects, mental health conditions (e.g., depression, anxiety), and socio-cultural influences on body image were often overlooked or inadequately adjusted in statistical analyses. Studies that did not stratify analyses to account for these confounders may have reported spurious associations between body image concerns and other psychological or health-related variables. For instance, studies that explored gender differences in body image dissatisfaction among PLWH often did not control for differential effects of ART-related side effects in men and women, leading to overgeneralized conclusions about gender-based disparities. Similarly, studies examining self-esteem and body dissatisfaction frequently lacked adjustments for psychosocial stressors (e.g., stigma, social support), which could be primary contributors to negative body image rather than biological or medical factors alone.

Additionally, the predominance of cross-sectional designs limits the ability to establish causal relationships between body image and its associated factors. Longitudinal studies are urgently needed to establish causal relationships between body image concerns and factors such as ART adherence, mental health outcomes, and social determinants to examine how body dissatisfaction evolves over time in response to ART side effects, changes in disease progression, and psychosocial influences.

The review also identified a lack of standardized measurement tools for assessing body image among PLWH. Sixteen different instruments were used across the studies, including MBSRQ, ABCD, and the Silhouette Scale. While these tools provided valuable insights, inconsistencies in their application limited the comparability of findings. Many studies failed to report the psychometric properties of their instruments or used adapted versions without validation, raising concerns about measurement reliability. Additionally, the lack of culturally tailored tools further reduced the applicability of findings across diverse populations. The reliance on self-reported BMI in a substantial portion of studies also highlights a key methodological limitation in body image research among PLWH. Given the known biases associated with self-reporting, future studies should prioritize clinician-measured BMI or, where feasible, incorporate advanced body composition assessments, which would offer a more precise evaluation of body composition, allowing for a more nuanced understanding of how BMI, fat distribution, and body image interact in this population.

Finally, the interplay between stigma, mental health, and body image in PLWH requires further investigation. Several studies in our review demonstrated that HIV-related stigma exacerbates body dissatisfaction and psychological distress, yet few studies examined stigma-reduction strategies as a means of improving body image outcomes. Only two studies provided an integrated intervention to address body image among PLWH, which underscores the effectiveness of this strategy in improving mental and physical outcomes in this population. However, a lack of follow-up and study settings in a single country/cultural context limited the generalizability of the findings.

There is a few interventions targeting body image issues among PLWH. Future research should explore alternative intervention approaches, including digital health interventions, community-based support programs, and integrated mental health services within HIV care. Also, studies should assess structural and policy-level interventions aimed at reducing stigma in healthcare and community settings, as well as the potential benefits of peer-led and group-based support models for PLWH struggling with body image concerns. Addressing these research gaps will be essential for developing comprehensive, culturally sensitive interventions that enhance the well-being and treatment outcomes of PLWH across diverse contexts.

### Our study limitations

A key strength of this review lies in its comprehensive synthesis of studies spanning over two decades, utilizing diverse methodologies, and considering international findings. This is the first review to comprehensively address the multidimensional aspects of body image concerns in PLWH. However, several limitations should be acknowledged. The decision to limit our search to only four databases may have restricted the scope of the literature review and potentially excluded relevant studies indexed in other databases. Additionally, the inclusion of only English-language, peer-reviewed studies, along with the exclusion of grey literature, may have led to the omission of valuable insights that could have further enriched the findings. For future literature reviews, we recommend the expansion of the search to include a wider range of databases and sources, including grey literature and non-English publications, to capture a more diverse set of studies. Moreover, incorporating studies from underrepresented geographic regions and targeting more interventional studies could enhance the generalizability and depth of findings on body image among PLWH.

### Clinical and Public Health Implications

The findings of this review underscore the critical need to integrate body image assessments into routine HIV care to identify and address body image-related distress among PLWH. Given the strong associations between body dissatisfaction, mental health conditions (e.g., depression, anxiety, BDD), and ART adherence, a holistic, multidisciplinary approach is essential. Incorporating regular screening for body image concerns within HIV care settings can facilitate the early identification of at-risk individuals, allowing for timely psychological and medical interventions.

Intervention strategies such as CBT-BISC have demonstrated effectiveness in reducing body dissatisfaction and improving ART adherence, yet their accessibility remains limited. To maximize reach and impact, integrating low-resource, scalable interventions, including digital mental health programs, peer-led support groups, and community-based psychosocial interventions, may offer more sustainable solutions, particularly in resource-limited settings. Furthermore, given the gender and racial disparities in body image concerns, interventions must be culturally adapted and tailored to meet the specific needs of women, youth, and racial/ethnic minorities living with HIV, who often experience heightened body dissatisfaction and stigma.

From a public health perspective, addressing structural barriers such as stigma, discrimination, and socioeconomic inequalities is crucial for improving body image outcomes. Health policies that promote stigma reduction, such as public awareness campaigns, healthcare provider training, and inclusive messaging around HIV-related body changes, could help reduce the psychosocial burden associated with body dissatisfaction. Moreover, integrating mental health services into HIV care frameworks will be essential for ensuring that body image interventions are not only available but also widely accessible and sustainable. By addressing body image concerns through a comprehensive, patient-centered approach, healthcare systems can enhance mental health outcomes, improve ART adherence, and ultimately contribute to better overall well-being for PLWH.

## Conclusion

Body image concerns among PLWH are a complex issue shaped by a combination of biopsychosocial factors. This review identified key risk factors, including HIV-related bodily changes (e.g., lipodystrophy, BMI fluctuations, ART side effects), psychological distress (e.g., depression, anxiety, BDD), and social determinants (e.g., stigma, gender bias). These concerns, in turn, influence ART adherence, mental health, and overall well-being. Protective factors such as strong social networks and targeted interventions like CBT-BISC have demonstrated potential in alleviating body image distress and improving self-perception among PLWH. Future research should focus on longitudinal studies, the development of validated body image measures for PLWH, and integrated healthcare approaches that incorporate both mental and physical health support. Addressing these research and clinical gaps will be essential for enhancing the well-being and quality of life of PLWH.

## Data Availability

All data produced in the present work are contained in the manuscript.

## Acknowledgements

We would like to acknowledge Irene Esu for her valuable contributions during the second-round search and screening process of this review study.

## Contribution

**Atena Pasha** (Conceptualization, Investigation, Methodology, Project administration, Resources, Writing – original draft, Writing – review and editing), **Mohammad Jahanaray** (Investigation, Data curation, Methodology, Formal analysis, Writing – original draft), **Xiaoming Li** (Supervision, Writing – review and editing), **Shan Qiao** (Supervision, Project administration, Methodology, Writing – review and editing).

---

**Figure 1.**
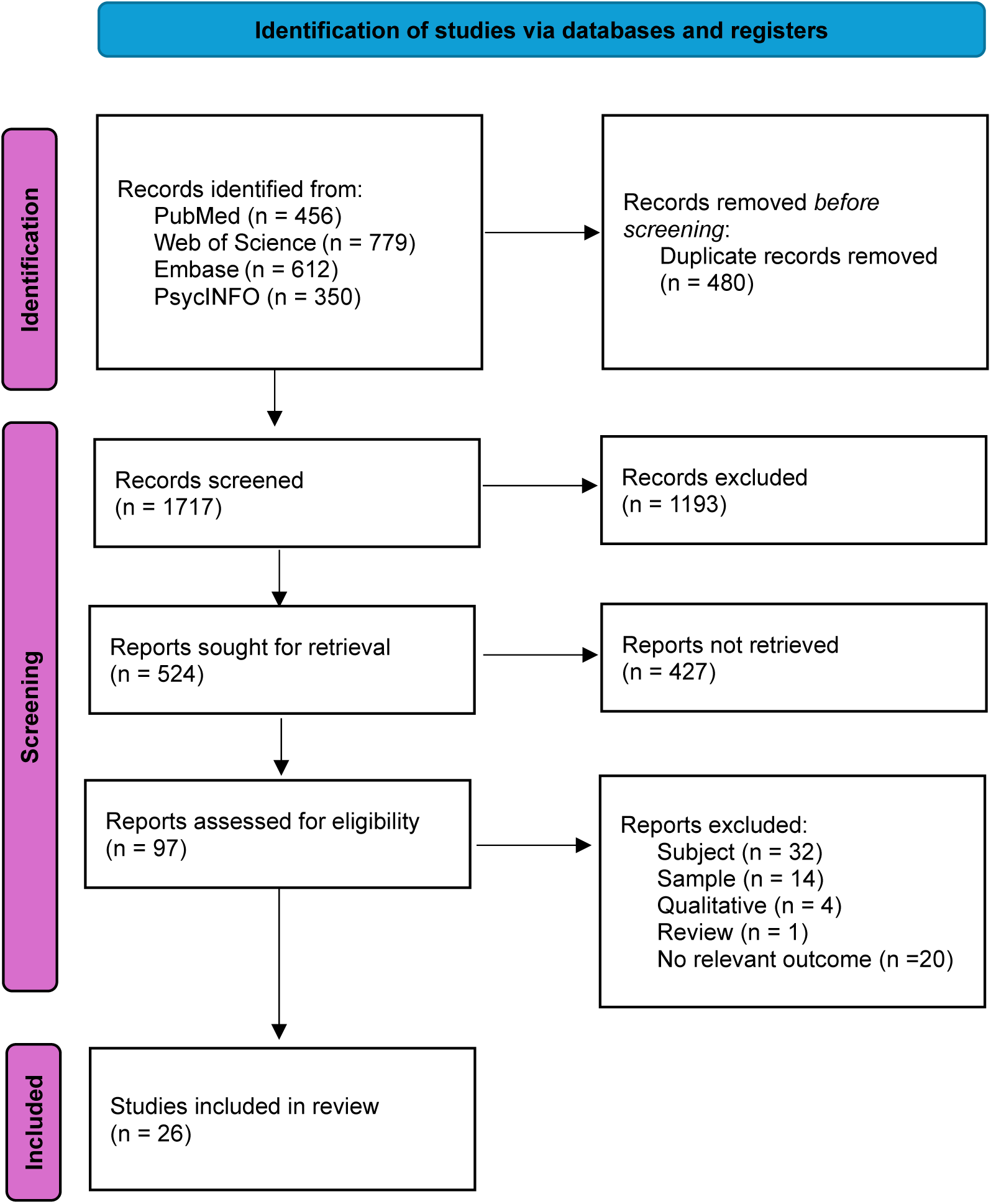
Flow diagram of the literature search and articles selection (adapted from PRISMA 2020 guidelines for systematic reviews).

## Footnotes

1 Difference in Pre-Post Change Scores, Squared

