## Supplemental document for "Body Image and Its Associated Factors among People Living with HIV: A Comprehensive Systematic Review and implications for integrated care"

**Supplementary Table 1a.** *Search strategy and keywords used for identifying studies related to people living with HIV and body image.*

| **Concept** | **Controlled Vocabulary** | **Keywords** |
| --- | --- | --- |
| PLWH | - "Acquired Immunodeficiency Syndrome"[Mesh] - "HIV"[Mesh] | - Persons living with AIDS - People living with AIDS - Persons living with HIV - People living with HIV - PLHIV - PLWH - PLWHA - Acquired immunodeficiency syndrome(s) - Acquired immune-deficiency syndrome(s) - Acquired immune deficiency syndrome(s) - AIDS - Human immunodeficiency virus(es) - HIV - HIV1 - HIV-1 - HIV2 - HIV-2 |
| Body Image | - “Body Image”[Mesh] - “Self-Concept”[Mesh:NoExp] | - Body image(s) - Body-image - Body integrity - Body schema(s) - Body representation(s) - Body dissatisfaction - Body satisfaction - Body esteem - Body self-esteem - Body appreciation - Body shame - Body preoccupation - Body discomfort - Body perception - Body insecurity - Body acceptance - Body confidence - Body concern - Body attitude - Body awareness - Body dysphoria - Body dysmorphia - Body dysmorphic - Body distortion - Body ideal - Self-image - Self-concept - Self-representation - Appearance evaluation - Appearance ~ (dis)satisfaction - Appearance anxiety - Appearance concern - Appearance-related anxiety - Appearance-related concern |

**Supplementary Table 1b.** *Search strategy and search string used in* ***PubMed****.*

| **Search #** | **Concept** | **Query** | **Results** |
| --- | --- | --- | --- |
| 1 | PLWH | "Acquired Immunodeficiency Syndrome"[Mesh] OR "HIV"[Mesh] OR “persons living with AIDS”[tiab] OR “people living with AIDS”[tiab] OR “persons living with HIV”[tiab] OR “people living with HIV”[tiab] OR PLHIV[tiab] OR PLWH[tiab] OR PLWHA[tiab] OR “acquired immunodeficiency syndrome*”[tiab] OR “acquired immune-deficiency syndrome*”[tiab] OR “acquired immune deficiency syndrome*”[tiab] OR AIDS[tiab] OR “human immunodeficiency virus*”[tiab] OR HIV[tiab] OR HIV1[tiab] OR HIV-1[tiab] OR HIV2[tiab] OR HIV-2[tiab] | 497,775 |
| 2 | Body Image | "Body Image"[Mesh] OR “body image*”[tiab] OR body-image[tiab] OR “body integrity”[tiab] OR “body schema*”[tiab] OR “body representation*”[tiab] OR “body dissatisfaction”[tiab] OR “body satisfaction”[tiab] OR “body esteem”[tiab] OR “body self-esteem”[tiab] OR “body appreciation”[tiab] OR “body shame”[tiab] OR “body preoccupation”[tiab] OR “body discomfort”[tiab] OR “body perception”[tiab] OR “body insecurity”[tiab] OR “body acceptance”[tiab] OR “body confidence”[tiab] OR “body concern*”[tiab] OR “body attitude*”[tiab] OR “body awareness”[tiab] OR “body dysphoria”[tiab] OR “body dysmorph*”[tiab] OR “body distortion”[tiab] OR “body ideal”[tiab] OR self-image[tiab] OR self-concept[tiab] OR self-representation[tiab] OR “appearance evaluation”[tiab] OR “appearance satisfaction”[tiab:~3] OR “appearance dissatisfaction”[tiab:~3] OR “appearance anxiety”[tiab] OR “appearance concern*”[tiab] OR “appearance-related anxiety”[tiab] OR “appearance-related concern*”[tiab] | 45,256 |
| 3 |  | #1 AND #2 | 533 |
| 4 | Language | #3 AND eng[la] | 511 |
| 5 | Date | #4 AND 2000/01/01:2024/12/31[pdat] | 456 |

("Acquired Immunodeficiency Syndrome"[Mesh] OR "HIV"[Mesh] OR "persons living with AIDS"[tiab] OR "people living with AIDS"[tiab] OR "persons living with HIV"[tiab] OR "people living with HIV"[tiab] OR PLHIV[tiab] OR PLWH[tiab] OR PLWHA[tiab] OR "acquired immunodeficiency syndrome*"[tiab] OR "acquired immune-deficiency syndrome*"[tiab] OR "acquired immune deficiency syndrome*"[tiab] OR AIDS[tiab] OR "human immunodeficiency virus*"[tiab] OR HIV[tiab] OR HIV1[tiab] OR HIV-1[tiab] OR HIV2[tiab] OR HIV-2[tiab]) AND ("Body Image"[Mesh] OR "body image*"[tiab] OR body-image[tiab] OR "body integrity"[tiab] OR "body schema*"[tiab] OR "body representation*"[tiab] OR "body dissatisfaction"[tiab] OR "body satisfaction"[tiab] OR "body esteem"[tiab] OR "body self-esteem"[tiab] OR "body appreciation"[tiab] OR "body shame"[tiab] OR "body preoccupation"[tiab] OR "body discomfort"[tiab] OR "body perception"[tiab] OR "body insecurity"[tiab] OR "body acceptance"[tiab] OR "body confidence"[tiab] OR "body concern*"[tiab] OR "body attitude*"[tiab] OR "body awareness"[tiab] OR "body dysphoria"[tiab] OR "body dysmorph*"[tiab] OR "body distortion"[tiab] OR "body ideal"[tiab] OR self-image[tiab] OR self-concept[tiab] OR self-representation[tiab] OR "appearance evaluation"[tiab] OR "appearance satisfaction"[tiab:~3] OR "appearance dissatisfaction"[tiab:~3] OR "appearance anxiety"[tiab] OR "appearance concern*"[tiab] OR "appearance-related anxiety"[tiab] OR "appearance-related concern*"[tiab]) AND eng[la] AND 2000/01/01:2024/12/31[pdat]

**Supplementary Table 1c.** *Search strategy and search string used in* ***PsycINFO****.*

| **Search #** | **Concept** | **Query** | **Results** |
| --- | --- | --- | --- |
| 1 | PLWH | ( DE "AIDS" OR DE "HIV" ) OR TI ( “persons living with AIDS” OR “people living with AIDS” OR “persons living with HIV” OR “people living with HIV” OR PLHIV OR PLWH OR PLWHA OR “acquired immunodeficiency syndrome*” OR “acquired immune-deficiency syndrome*” OR “acquired immune deficiency syndrome*” OR AIDS OR “human immunodeficiency virus*” OR HIV OR HIV1 OR HIV-1 OR HIV2 OR HIV-2 ) OR AB ( “persons living with AIDS” OR “people living with AIDS” OR “persons living with HIV” OR “people living with HIV” OR PLHIV OR PLWH OR PLWHA OR “acquired immunodeficiency syndrome*” OR “acquired immune-deficiency syndrome*” OR “acquired immune deficiency syndrome*” OR AIDS OR “human immunodeficiency virus*” OR HIV OR HIV1 OR HIV-1 OR HIV2 OR HIV-2 ) | 79,703 |
| 2 | Body Image | ( DE "Body Image" OR DE "Body Dissatisfaction" OR DE "Body Esteem") OR TI ( “body image*” OR body-image OR “body integrity” OR “body schema*” OR “body representation*” OR “body dissatisfaction” OR “body satisfaction” OR “body esteem” OR “body self-esteem” OR “body appreciation” OR “body shame” OR “body preoccupation” OR “body discomfort” OR “body perception” OR “body insecurity” OR “body acceptance” OR “body confidence” OR “body concern*” OR “body attitude*” OR “body awareness” OR “body dysphoria” OR “body dysmorph*” OR “body distortion” OR “body ideal” OR self-image OR self-concept OR self-representation OR “appearance evaluation” OR (appearance N3 satisfaction) OR (appearance N3 dissatisfaction) OR “appearance anxiety” OR “appearance concern*” OR “appearance-related anxiety” OR “appearance-related concern*” ) OR AB ( “body image*” OR body-image OR “body integrity” OR “body schema*” OR “body representation*” OR “body dissatisfaction” OR “body satisfaction” OR “body esteem” OR “body self-esteem” OR “body appreciation” OR “body shame” OR “body preoccupation” OR “body discomfort” OR “body perception” OR “body insecurity” OR “body acceptance” OR “body confidence” OR “body concern*” OR “body attitude*” OR “body awareness” OR “body dysphoria” OR “body dysmorph*” OR “body distortion” OR “body ideal” OR self-image OR self-concept OR self-representation OR “appearance evaluation” OR (appearance N3 satisfaction) OR (appearance N3 dissatisfaction) OR “appearance anxiety” OR “appearance concern*” OR “appearance-related anxiety” OR “appearance-related concern*” ) | 60,382 |
| 3 |  | #1 AND #2 | 476 |
| 4 | Language | #3 AND LA English | 445 |
| 5 | Date | #4 AND PY 2000-2024 | 350 |

( ( DE "AIDS" OR DE "HIV" ) OR TI ( “persons living with AIDS” OR “people living with AIDS” OR “persons living with HIV” OR “people living with HIV” OR PLHIV OR PLWH OR PLWHA OR “acquired immunodeficiency syndrome*” OR “acquired immune-deficiency syndrome*” OR “acquired immune deficiency syndrome*” OR AIDS OR “human immunodeficiency virus*” OR HIV OR HIV1 OR HIV-1 OR HIV2 OR HIV-2 ) OR AB ( “persons living with AIDS” OR “people living with AIDS” OR “persons living with HIV” OR “people living with HIV” OR PLHIV OR PLWH OR PLWHA OR “acquired immunodeficiency syndrome*” OR “acquired immune-deficiency syndrome*” OR “acquired immune deficiency syndrome*” OR AIDS OR “human immunodeficiency virus*” OR HIV OR HIV1 OR HIV-1 OR HIV2 OR HIV-2 ) ) AND ( ( DE "Body Image" OR DE "Body Dissatisfaction" OR DE "Body Esteem") OR TI ( “body image*” OR body-image OR “body integrity” OR “body schema*” OR “body representation*” OR “body dissatisfaction” OR “body satisfaction” OR “body esteem” OR “body self-esteem” OR “body appreciation” OR “body shame” OR “body preoccupation” OR “body discomfort” OR “body perception” OR “body insecurity” OR “body acceptance” OR “body confidence” OR “body concern*” OR “body attitude*” OR “body awareness” OR “body dysphoria” OR “body dysmorph*” OR “body distortion” OR “body ideal” OR self-image OR self-concept OR self-representation OR “appearance evaluation” OR (appearance N3 satisfaction) OR (appearance N3 dissatisfaction) OR “appearance anxiety” OR “appearance concern*” OR “appearance-related anxiety” OR “appearance-related concern*” ) OR AB ( “body image*” OR body-image OR “body integrity” OR “body schema*” OR “body representation*” OR “body dissatisfaction” OR “body satisfaction” OR “body esteem” OR “body self-esteem” OR “body appreciation” OR “body shame” OR “body preoccupation” OR “body discomfort” OR “body perception” OR “body insecurity” OR “body acceptance” OR “body confidence” OR “body concern*” OR “body attitude*” OR “body awareness” OR “body dysphoria” OR “body dysmorph*” OR “body distortion” OR “body ideal” OR self-image OR self-concept OR self-representation OR “appearance evaluation” OR (appearance N3 satisfaction) OR (appearance N3 dissatisfaction) OR “appearance anxiety” OR “appearance concern*” OR “appearance-related anxiety” OR “appearance-related concern*” ) ) AND LA English AND PY 2000-2024

**Supplementary Table 1d.** *Search strategy and search string used in* ***Embase****.*

| **Search #** | **Concept** | **Query** | **Results** |
| --- | --- | --- | --- |
| 1 | PLWH | 'aids patient'/exp OR 'human immunodeficiency virus infected patient'/exp OR 'acquired immune deficiency syndrome'/de OR 'human immunodeficiency virus'/exp OR 'persons living with aids':ab,ti OR 'people living with aids':ab,ti OR 'persons living with hiv':ab,ti OR 'people living with hiv':ab,ti OR plhiv:ab,ti OR plwh:ab,ti OR plwha:ab,ti OR 'acquired immunodeficiency syndrome*':ab,ti OR 'acquired immune-deficiency syndrome*':ab,ti OR 'acquired immune deficiency syndrome*':ab,ti OR aids:ab,ti OR 'human immunodeficiency virus*':ab,ti OR hiv:ab,ti OR hiv1:ab,ti OR 'hiv 1':ab,ti OR hiv2:ab,ti OR 'hiv 2':ab,ti | 661,086 |
| 2 | Body Image | 'body image'/exp OR 'body representation'/exp OR 'body satisfaction'/exp OR 'body esteem'/exp OR 'body appreciation'/exp OR 'body image*':ab,ti OR 'body integrity':ab,ti OR 'body schema*':ab,ti OR 'body representation*':ab,ti OR 'body dissatisfaction':ab,ti OR 'body satisfaction':ab,ti OR 'body esteem':ab,ti OR 'body self-esteem':ab,ti OR 'body appreciation':ab,ti OR 'body shame':ab,ti OR 'body preoccupation':ab,ti OR 'body discomfort':ab,ti OR 'body perception':ab,ti OR 'body insecurity':ab,ti OR 'body acceptance':ab,ti OR 'body confidence':ab,ti OR 'body concern*':ab,ti OR 'body attitude*':ab,ti OR 'body awareness':ab,ti OR 'body dysphoria':ab,ti OR 'body dysmorph*':ab,ti OR 'body distortion':ab,ti OR 'body ideal':ab,ti OR 'self image':ab,ti OR 'self concept':ab,ti OR 'self representation':ab,ti OR 'appearance evaluation':ab,ti OR (appearance NEAR/3 satisfaction) OR (appearance NEAR/3 dissatisfaction) OR 'appearance anxiety':ab,ti OR 'appearance concern*':ab,ti OR 'appearance-related anxiety':ab,ti OR 'appearance-related concern*':ab,ti | 54,545 |
| 3 |  | #1 AND #2 | 689 |
| 4 | Language | #3 AND [english]/lim | 663 |
| 5 | Date | #4 AND [2000-2024]/py | 612 |

('aids patient'/exp OR 'human immunodeficiency virus infected patient'/exp OR 'acquired immune deficiency syndrome'/de OR 'human immunodeficiency virus'/exp OR 'persons living with aids':ab,ti OR 'people living with aids':ab,ti OR 'persons living with hiv':ab,ti OR 'people living with hiv':ab,ti OR plhiv:ab,ti OR plwh:ab,ti OR plwha:ab,ti OR 'acquired immunodeficiency syndrome*':ab,ti OR 'acquired immune-deficiency syndrome*':ab,ti OR 'acquired immune deficiency syndrome*':ab,ti OR aids:ab,ti OR 'human immunodeficiency virus*':ab,ti OR hiv:ab,ti OR hiv1:ab,ti OR 'hiv 1':ab,ti OR hiv2:ab,ti OR 'hiv 2':ab,ti) AND ('body image'/exp OR 'body representation'/exp OR 'body satisfaction'/exp OR 'body esteem'/exp OR 'body appreciation'/exp OR 'body image*':ab,ti OR 'body integrity':ab,ti OR 'body schema*':ab,ti OR 'body representation*':ab,ti OR 'body dissatisfaction':ab,ti OR 'body satisfaction':ab,ti OR 'body esteem':ab,ti OR 'body self-esteem':ab,ti OR 'body appreciation':ab,ti OR 'body shame':ab,ti OR 'body preoccupation':ab,ti OR 'body discomfort':ab,ti OR 'body perception':ab,ti OR 'body insecurity':ab,ti OR 'body acceptance':ab,ti OR 'body confidence':ab,ti OR 'body concern*':ab,ti OR 'body attitude*':ab,ti OR 'body awareness':ab,ti OR 'body dysphoria':ab,ti OR 'body dysmorph*':ab,ti OR 'body distortion':ab,ti OR 'body ideal':ab,ti OR 'self image':ab,ti OR 'self concept':ab,ti OR 'self representation':ab,ti OR 'appearance evaluation':ab,ti OR (appearance NEAR/3 satisfaction) OR (appearance NEAR/3 dissatisfaction) OR 'appearance anxiety':ab,ti OR 'appearance concern*':ab,ti OR 'appearance-related anxiety':ab,ti OR 'appearance-related concern*':ab,ti) AND [english]/lim AND [2000-2024]/py

**Supplementary Table 1e.** *Search strategy and search string used in* ***Web of Science****.*

| **Search #** | **Concept** | **Query** | **Results** |
| --- | --- | --- | --- |
| 1 | PLWH | (TI=(“persons living with AIDS” OR “people living with AIDS” OR “persons living with HIV” OR “people living with HIV” OR PLHIV OR PLWH OR PLWHA OR “acquired immunodeficiency syndrome*” OR “acquired immune-deficiency syndrome*” OR “acquired immune deficiency syndrome*” OR AIDS OR “human immunodeficiency virus*” OR HIV OR HIV1 OR HIV-1 OR HIV2 OR HIV-2 )) OR AB=(“persons living with AIDS” OR “people living with AIDS” OR “persons living with HIV” OR “people living with HIV” OR PLHIV OR PLWH OR PLWHA OR “acquired immunodeficiency syndrome*” OR “acquired immune-deficiency syndrome*” OR “acquired immune deficiency syndrome*” OR AIDS OR “human immunodeficiency virus*” OR HIV OR HIV1 OR HIV-1 OR HIV2 OR HIV-2 ) | 1,077,196 |
| 2 | Body Image | (TI=(“body image*” OR body-image OR “body integrity” OR “body schema*” OR “body representation*” OR “body dissatisfaction” OR “body satisfaction” OR “body esteem” OR “body self-esteem” OR “body appreciation” OR “body shame” OR “body preoccupation” OR “body discomfort” OR “body perception” OR “body insecurity” OR “body acceptance” OR “body confidence” OR “body concern*” OR “body attitude*” OR “body awareness” OR “body dysphoria” OR “body dysmorph*” OR “body distortion” OR “body ideal” OR self-image OR self-concept OR self-representation OR “appearance evaluation” OR (appearance NEAR/3 satisfaction) OR (appearance NEAR/3 dissatisfaction) OR “appearance anxiety” OR “appearance concern*” OR “appearance-related anxiety” OR “appearance-related concern*”)) OR AB=(“body image*” OR body-image OR “body integrity” OR “body schema*” OR “body representation*” OR “body dissatisfaction” OR “body satisfaction” OR “body esteem” OR “body self-esteem” OR “body appreciation” OR “body shame” OR “body preoccupation” OR “body discomfort” OR “body perception” OR “body insecurity” OR “body acceptance” OR “body confidence” OR “body concern*” OR “body attitude*” OR “body awareness” OR “body dysphoria” OR “body dysmorph*” OR “body distortion” OR “body ideal” OR self-image OR self-concept OR self-representation OR “appearance evaluation” OR (appearance NEAR/3 satisfaction) OR (appearance NEAR/3 dissatisfaction) OR “appearance anxiety” OR “appearance concern*” OR “appearance-related anxiety” OR “appearance-related concern*”) | 58,589 |
| 3 |  | #1 AND #2 | 879 |
| 4 | Language | #3 AND (LA=English) | 841 |
| 5 | Date | #4 AND (PY=(2000-2024)) | 779 |

((TI=(“persons living with AIDS” OR “people living with AIDS” OR “persons living with HIV” OR “people living with HIV” OR PLHIV OR PLWH OR PLWHA OR “acquired immunodeficiency syndrome*” OR “acquired immune-deficiency syndrome*” OR “acquired immune deficiency syndrome*” OR AIDS OR “human immunodeficiency virus*” OR HIV OR HIV1 OR HIV-1 OR HIV2 OR HIV-2 )) OR AB=(“persons living with AIDS” OR “people living with AIDS” OR “persons living with HIV” OR “people living with HIV” OR PLHIV OR PLWH OR PLWHA OR “acquired immunodeficiency syndrome*” OR “acquired immune-deficiency syndrome*” OR “acquired immune deficiency syndrome*” OR AIDS OR “human immunodeficiency virus*” OR HIV OR HIV1 OR HIV-1 OR HIV2 OR HIV-2 )) AND ((TI=(“body image*” OR body-image OR “body integrity” OR “body schema*” OR “body representation*” OR “body dissatisfaction” OR “body satisfaction” OR “body esteem” OR “body self-esteem” OR “body appreciation” OR “body shame” OR “body preoccupation” OR “body discomfort” OR “body perception” OR “body insecurity” OR “body acceptance” OR “body confidence” OR “body concern*” OR “body attitude*” OR “body awareness” OR “body dysphoria” OR “body dysmorph*” OR “body distortion” OR “body ideal” OR self-image OR self-concept OR self-representation OR “appearance evaluation” OR (appearance NEAR/3 satisfaction) OR (appearance NEAR/3 dissatisfaction) OR “appearance anxiety” OR “appearance concern*” OR “appearance-related anxiety” OR “appearance-related concern*”)) OR AB=(“body image*” OR body-image OR “body integrity” OR “body schema*” OR “body representation*” OR “body dissatisfaction” OR “body satisfaction” OR “body esteem” OR “body self-esteem” OR “body appreciation” OR “body shame” OR “body preoccupation” OR “body discomfort” OR “body perception” OR “body insecurity” OR “body acceptance” OR “body confidence” OR “body concern*” OR “body attitude*” OR “body awareness” OR “body dysphoria” OR “body dysmorph*” OR “body distortion” OR “body ideal” OR self-image OR self-concept OR self-representation OR “appearance evaluation” OR (appearance NEAR/3 satisfaction) OR (appearance NEAR/3 dissatisfaction) OR “appearance anxiety” OR “appearance concern*” OR “appearance-related anxiety” OR “appearance-related concern*”)) AND (LA=English) AND (PY=(2000-2024))
